# Metagenomic analysis of colonic tissue and stool microbiome in patients with colorectal cancer in a South Asian population

**DOI:** 10.1101/2024.01.25.24301775

**Authors:** Bawantha Dilshan Gamage, Diyanath Ranasinghe, AGP Sahankumari, Gathsaurie Neelika Malavige

## Abstract

**Objective:** The gut microbiome is thought to play an important role in the development of colorectal cancer (CRC). However, as the gut microbiome varies widely based on diet, we sought to investigate the gut microbiome changes in patients with CRC in a South Asian population.

**Design:** The gut microbiome was assessed by 16s metagenomic sequencing targeting the V4 hypervariable region of the bacterial 16S rRNA in stool samples (n=112) and colonic tissue (n=36) in 112 individuals. Of these had CRC (n=24), premalignant lesions (n=10), healthy individuals (n=50) and in those with diabetes (n=28).

**Results:** Overall, the relative abundances of genus Fusobacterium (p < 0.001), Acinetobacter (p < 0.001), Escherichia-Shigella (p < 0.05) were significantly higher in gut tissue, while Romboutsia (p < 0.01) and Prevotella (p < 0.05) were significantly higher in stool samples. Bacteroides and Fusobacterium, were the most abundant genera found in stool samples in patients with CRC. Patients with pre-malignant lesions had significantly high abundances of Christensenellaceae, Enterobacteriaceae, Mollicutes and Ruminococcaceae (p < 0.001) compared to patients with CRC, and healthy individuals. Romboutsia was significantly more abundant (p< 0.01) in stool samples in healthy individuals compared to those with CRC and diabetes.

**Conclusion:** Despite marked differences in the Sri Lankan diet compared to the typical Western diet, Bacteroides and Fusobacterium species were the most abundant in those with CRC, with *Prevotella* species, being most abundant in many individuals. We believe these results pave the way for possible dietary interventions for prevention of CRC in the South Asian population.

## Introduction

Colorectal cancers (CRC) are the third leading cancer, with approximately 1.9 million new cases and 930,000 deaths reported globally in 2020. With the increase in its prevalence, the deaths are predicted to increase to 1.6 million by 2040^1^. Although South Asia has reported a lower age standardized incidence of 14.8/100,000 person-years for males and 18.4/100,000 person years for females in 2020 compared to high income countries, the prevalence is gradually increasing^1^. In Sri Lanka, the age standardized incidence was 10.2/100,000 person years in 2019^2^ and the prevalence is likely to increase due to the change in population demographics, diet and lifestyle changes as seen in many countries in South Asia.

Risk factors for the development of CRC have been identified and include smoking, inflammatory bowel disease, diet, obesity, lack of exercise and alcohol consumption^3^. Dietary patterns such as consumption of red meat, a diet low in calcium and fiber and a diet low in milk have shown to be risk factors for development of CRC^3^. Obesity and hyperglycemia are also well-defined strong risk factors^4^. The increased prevalence of diabetes ^5^, obesity along with the change in diet is likely to lead to a significant increase in CRC, in regions such as South Asia, where currently a lower incidence is reported. While the mechanisms by which diet, obesity and diabetes contribute to development of CRC is not known, these directly influence the composition and diversity of the gut microbiome^6^ ^7^. CRC is associated with gut microbial dysbiosis with overabundance of bacteria such as *Bacteroides fragilis*, *Escherichia coli*, *Enterococcus faecalis*, *Fusobacterium nucleatum* and *Streptococcus gallolyticus*, with *Bacteroides fragilis* being the commonest bacterial strain^8^ ^9^. In fact, *Bacteroides fragilis* along with species such as *Fusobacterium nucleatum, Porphyromonas asaccharolytica, Parvimonas micra*, *Prevotella intermedia*, were shown to be overabundant in patients with CRC across many geographical locations such as Europe, America and China^10^. Higher abundance of Fusobacterium species has also been reported in adenomas, which are premalignant lesions leading to CRC^9^.

The diet is an important factor that leads to gut microbial diversity, and low intake of milk, fiber and whole grain products along with high intake of red meat being significantly associated with the risk of developing CRC^11^. The diet is known to play a significant role in the composition of the gut microbiome, with temporality changes occurring even within 24 hours of consuming a different diet^12^. Prolonged dietary changes are known to induce prolonged and permanent changes in the gut microbiome^13^. The gut microbiome shows markedly diversity based on the geographical location, with significant differences in Westen populations compared to African and South American populations^14^. In India, individuals from regions primarily consuming a plant-based diet were found to have an overabundance of Prevotella species compared to those who were consuming animal and plant-based products, who had abundance of the species Bacteroides, Ruminococcus, and Faecalibacterium^15^. Therefore, there are significant differences in the gut microbiome of the South Asian population compared to other populations, which may vary in those with CRC.

Although species such as *Bacteroides fragilis*, *Fusobacterium nucleatum*, *Porphyromonas asaccharolytica* etc. have shown to associate with CRC in the West and in Chinese communities where a red meat is readily consumed, red meat consumption is very much less in South Asian countries such as India and Sri Lanka. Therefore, the type of bacterial phyla and genera associated with CRC could differ. Furthermore, except for a few studies in India, there are very few studies describing the composition of the gut microbiome in the South Asian population, while there are none from Sri Lanka. In this study, we evaluated the gut microbiome in health individuals, individuals with CRC, individuals with diabetes did not have CRC or premalignant lesions (age and sex matched) and in those with premalignant lesions. This data would be important to identify suitable dietary interventions to possibly change the gut microbiome composition, as potential prevention strategies for development of CRC.

## Methods

### Participant characteristics

We recruited 112 individuals who underwent colonoscopy at Colombo South Teaching Hospital, Sri Lanka between January 2017, and April 2018, following informed written consent. All clinical details regarding altered bowel habits, abdominal pain, loss of weight, appetite along with laboratory and radiological investigations such as full blood count, ultrasound scanning of the abdomen and CT scans were recorded. Biopsies were obtained at the time of colonoscopy from the rectum, hepatic flexure, transverse colon, sigmoid colon and anal verge, as a part of their routine colonoscopy. The biopsies were then evaluated for the presence of CRC and grading was carried out according to TNM staging classification. The clinical characteristics of these individuals are shown in supplementary table 1.

### Collection of stool and biopsy samples used for microbiome studies

Stool samples were collected 2 weeks following colonoscopy from the above cohort and included 24 patients who were confirmed as having CRC, 10 with those who were found to have premalignant lesions, 50 healthy individuals (normal colonoscopy who did not have diabetes or any other illnesses) and 28 patients with diabetes mellitus (who had normal colonoscopy findings). The fasting blood sugar and lipid profiles were assessed in the 50 healthy individuals to exclude the presence of diabetes and hyperlipidemia. Biopsies of 18 patients who were confirmed to have CRC and biopsies of 18 healthy individuals were also included in the analysis of the microbiome.

### DNA extraction and Metagenomics analysis

The biopsy samples and stool samples were transported to the laboratory within 24 hours after collection. DNA from stool samples was extracted using DNeasy Powersoil kit (QS, Hilden, Germany) whereas DNA from tissue samples was extracted using DNeasy PowerLyzer Tissue and Cells Kit (QS, Hilden, Germany) according to the manufacturer’s instructions. Extracted DNA was stored at -80 C until sequencing was carried out. 16s metagenomic sequencing was carried out by Diversigen, USA. The V4 hypervariable region of the bacterial 16S rRNA marker gene (16Sv4) was PCR-amplified in duplicate with primers 515F-OH1 and 806R-OH2, which enabled us to characterize the gut microbiome up to the genus level. DNA libraries were prepared using PCR products according to the Nextera XT DNA Library Preparation kit guide (Illumina, CA, USA). These were then pooled and sequenced on the Illumina MiSeq platform with a read length of 2x250bp.

### Bioinformatics Analysis

Raw pyrosequencing read pairs were de-multiplexed according to the unique molecular barcodes using the MiSeq (Illumina) inbuilt tools. Resulting FASTQ reads were processed with the USEARCH ^16^ suite adhering to the best practices of the USEARCH guidelines. Paired-end reads were merged via fastq_mergepairs command with a minimum overlap of 50 bases and maximum mismatches of 5. USEARCH quality filter was set to discard merged reads containing above 5% mismatches. Singletons (unique sequences that are found only once), were discarded using USEARCH command as they can create many spurious OTUs during the downstream analysis. Also, chimeric sequences were detected and removed with UCHIME (Edgar, 2016). Clustering of the merged sequences into operational taxonomic units (OTUs) was performed using UPARSE-OTU algorithm ^16^. Then the sequences were binned at a similarity threshold of 97% and a list of representative OTU sequences was generated. Diversity and taxonomic analysis were done by mapping the representative OTUs against the SILVA, version 132 16S database ^17^. The OTU abundance table and the taxonomy table were created with USEARCH commands while a 16S rRNA gene-based phylogenetic tree was generated from representative FASTA sequences. Beta diversity matrixes were generated using the phylogenetic tree, which was plotted during the statistical analysis. OTU counts were normalized across the samples to 2965, which was the lowest number of reads a sample had acquired, to avoid potential bias caused by differing sequencing depths.

A total of 2,513,198 raw sequencing read pairs were obtained with a median count of 17,375 (SD = 6202) read pairs per sample. After filtering low-quality reads, artifacts, and singletons, 1,786,919 pairs (71.1%) were mapped against the 16S database. The highest mapped read count in a sample was 19,349 while the lowest being 2965 reads, hence all the samples were rarefied to 2965 reads.

### Statistical Analysis

The resulting OTU table, taxonomy table and sample data tables were analyzed with Rstudio\R version 4.0.3 ^18^ using the phyloseq package, version 3.12 ^19^ to generate abundance data for each hierarchical level. Alpha diversity in bacterial communities was calculated by R vegan package, version 2.4 ^20^ and presented by the number of observed OTUs and Shannon index. Non-parametric two-sample t-test was used to compare the alpha diversity metrics between the healthy and CRC samples. Weighted and unweighted UniFrac distances of beta diversity was plotted with principal coordinates analysis (PCoA). Analysis of similarity between gut tissue and stool samples were calculated with Bray-Curtis dissimilarity model using the “anosim” function at 1,000 permutations and species accumulation curves with the “specaccum” function in vegan package. All visualizations were done using R\ggplot2.

OTU counts were converted to relative abundance percentages for each sample using the R\funrar, version 1.4.1 ^21^. Significant differences between the healthy and CRC groups for gut tissue samples were calculated at phyla and genera hierarchical levels using the Mann–Whitney U test. For stool samples of healthy vs those with CRC, healthy vs those with a pre-malignant lesion and healthy vs diabetes, the groups were tested at phyla and genera levels using Kruskal-Wallis test. P values were corrected using Benjamini-Hochberg false-discovery rate (FDR) ^22^ and the significance assessed at 0.05. As an additional step, to compare the microbiome of the cohort of patients with diabetes with that of the groups who were classified as having pre-malignant lesions or and CRC, Kruskal-Wallis test with FDR corrections using Benjamini-Hochberg was used for analysis at phyla and genera levels. This was followed by a post-hoc analysis using Dunn’s multiple comparisons test on taxa that were identified to be different to determine which pair/pairs of groups were different among the subgroups.

## Results

### Overall comparison of the faecal and tissue microbiome in healthy individuals and patients with CRC

We assessed the gut microbiome in a total of 148 samples, of which 112 were stool samples and 36 tissue biopsy samples. The resulting OTU abundance data consisted of 818 bacterial genera belonging to 17 different phyla. Species accumulation curves (Figure 1) of each group against sampling effort (sites) indicated the highest species discovery in healthy stool samples. However, all the curves appear to overlap, except the curve for healthy gut tissue samples that exhibited slightly less species discovery over sampling effort.

**Figure 1:**
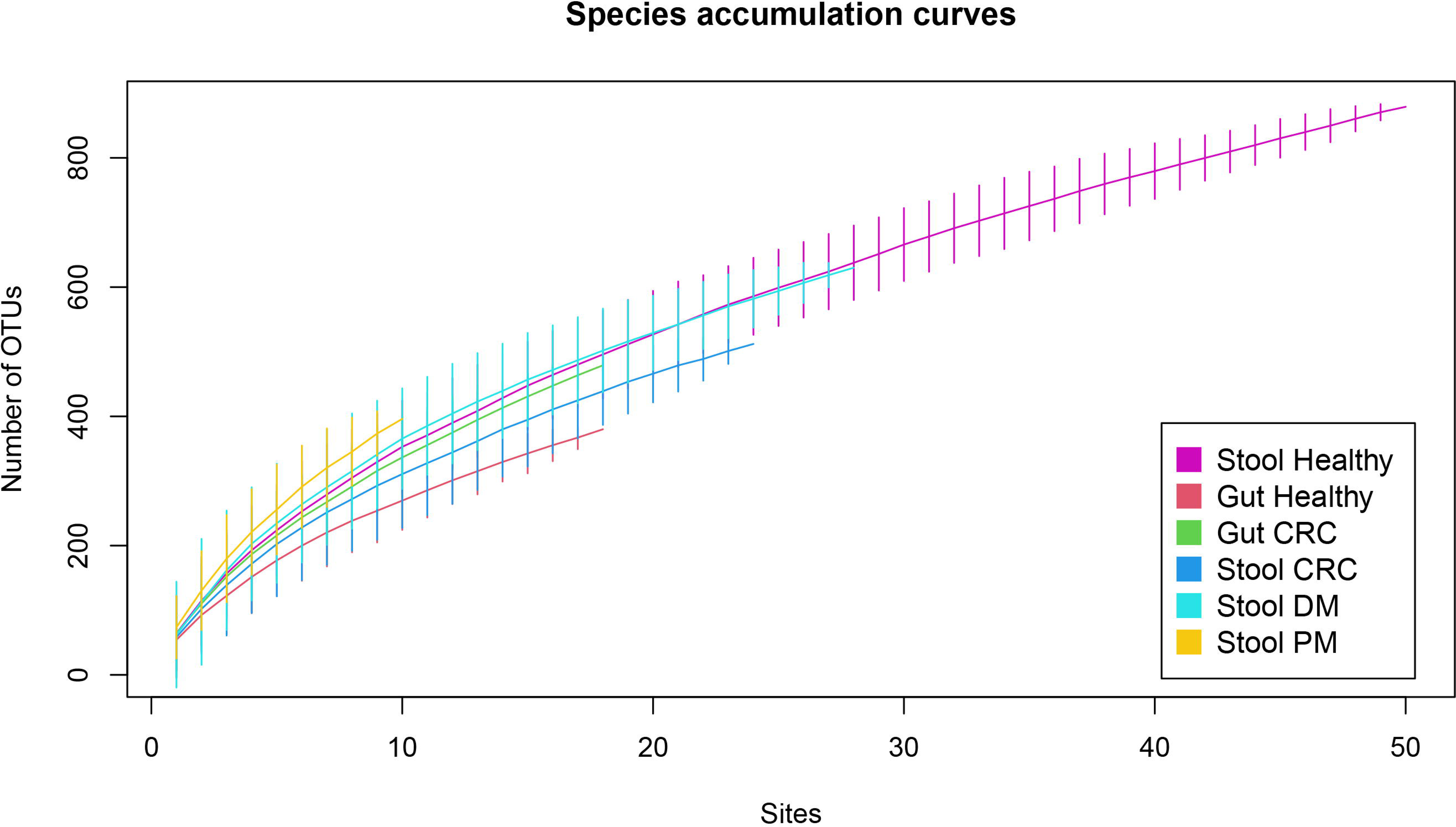
Species accumulation curves of healthy and CRC patients for each sample type. Healthy stool samples demonstrated the highest OTU discovery, whereas healthy gut tissue samples showed the least OTU discovery.

Microbial diversities between and within the sampling sites and their subgroups were measured using Shannon and Simpson indexes. In general, tissue biopsies showed higher richness and evenness suggesting higher bacterial complexity compared to faecal samples, although these differences were not significant. Also, there were no significant differences between the stool and tissue subgroups (Figure 2). However, beta diversity between stool and tissue samples was tested by mapping weighted UniFrac distances into Principal Coordinates Analysis (PCoA), which indicated significant clustering between the microbial composition of gut and stool samples (R = 0.0507, p = 0.001, Figure 3A). Although no differences were found in the microbial diversity between stools and tissues, there were significant differences in stool samples between the different patient sub-groups (healthy, those with premalignant lesions and those with CRC) when analyzed using the anosim (R = 0.121, P < 0.001) (Figure 3B).

**Figure 2:**
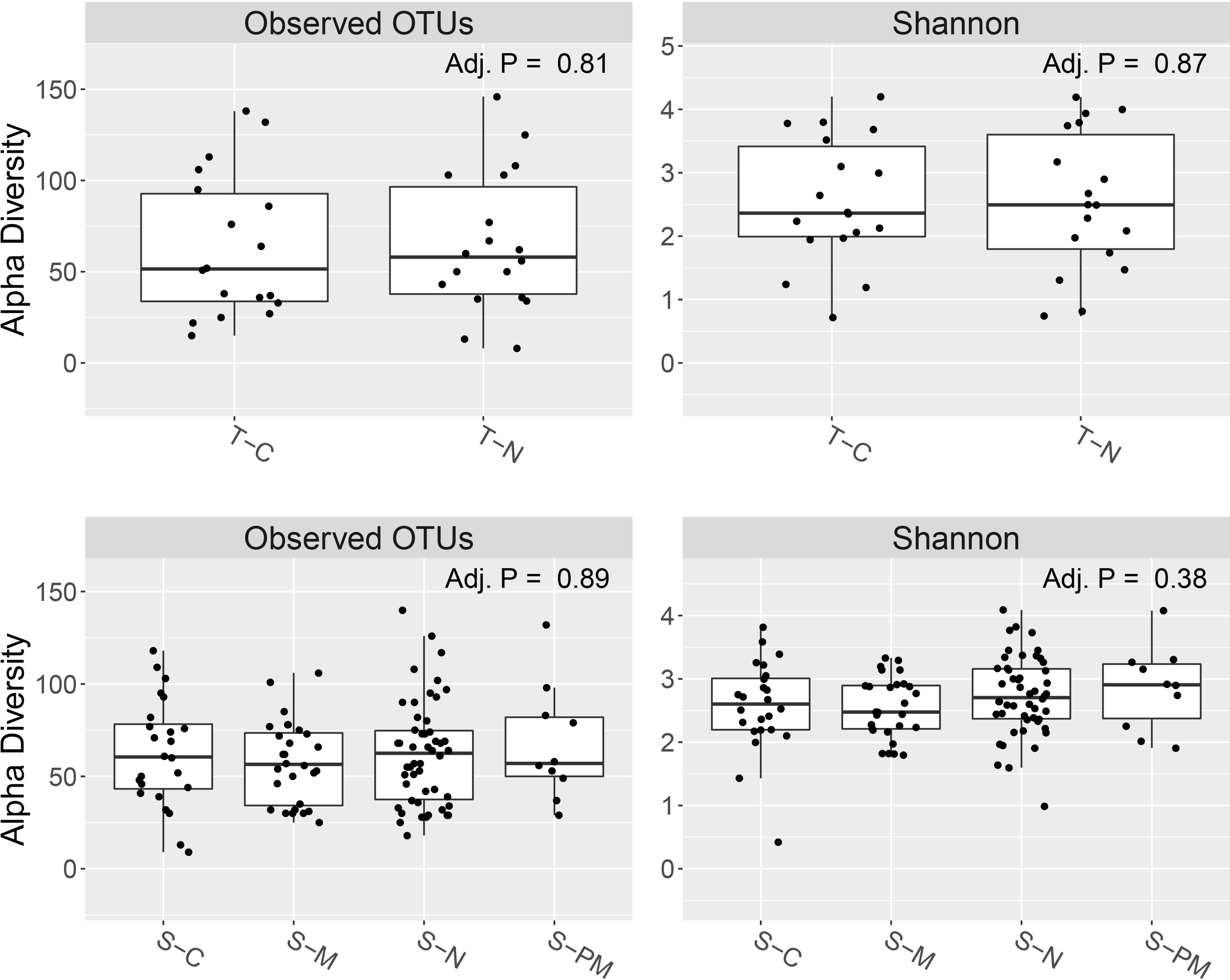
Comparison of the faecal and tissue microbiome (alpha diversity) in healthy individuals, patients with CRC, with premalignant lesions and with diabetes. Alpha diversity in bacterial communities was calculated and presented by the number of observed OTUs and Shannon index in tissue samples (T) and stool samples (S), in the in those with CRC (C), healthy individuals (N), those with diabetes (M) and in those with premalignant lesions (PM). P values were calculated with Kruskal-Wallis test.

**Figure 3:**
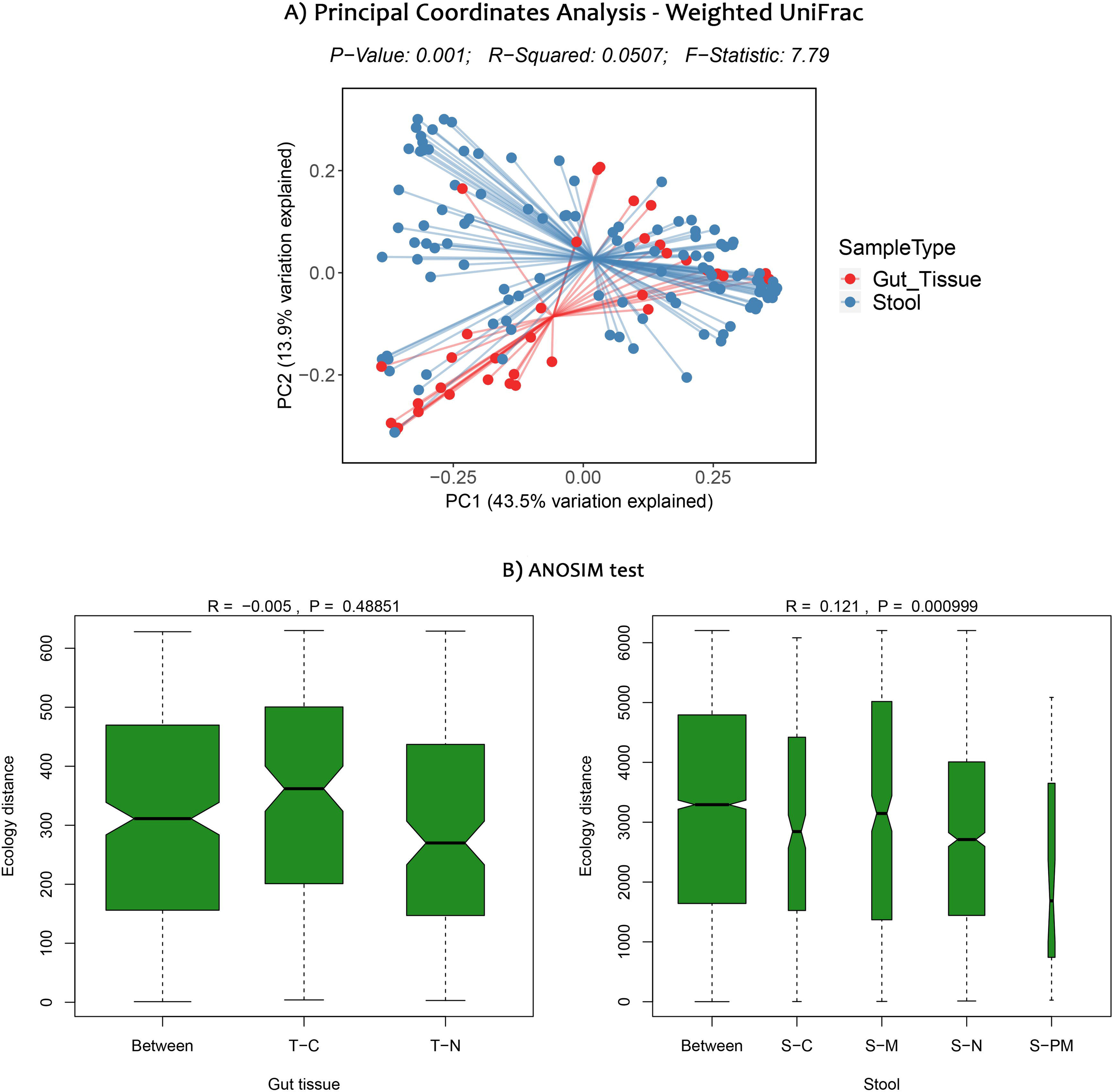
Comparison of the beta diversity of faecal and tissue microbiome in healthy individuals, patients with CRC, with premalignant lesions and with diabetes. The similarity test (ANOSIM) for samples obtained from colonic tissue (T) and stools (S), in those with CRC (C), healthy individuals (N), those with diabetes (M) and in those with premalignant lesions (PM) was carried out to assess the differences in microbial composition. The microbial composition between gut tissue subgroups was consistent while stool samples exhibit significant dissimilarity (p < 0.0001) between subgroups (A). The Principal Coordinates Analysis (PCoA) using weighted UniFrac distances indicating significant clustering between two types of samples, gut tissue and stool, (*p* = 0.001) (B).

### Compositional analysis of the stool and tissue samples of patients and healthy individuals

The overall analysis of the taxonomic composition in all subgroups (health, those with CRC, premalignant lesions and diabetes) revealed that more than 97% of the sequences collected were classified into six dominant phyla: Firmicutes (37%), Bacteroidetes (31%), Proteobacteria (19%), Actinobacteria (7%), Fusobacteria (2%) and Verrucomicrobia (2%). However, the relative abundances of Fusobacteria (p < 0.001), and Proteobacteria (p < 0.005) were overall significantly higher in tissue samples compared to stool samples, whereas Firmicutes (p = 0.03) and Actinobacteria (p = 0.03) were significantly abundant in stool samples. At genera level, overall relative abundances of genus Fusobacterium (p < 0.001), Acinetobacter (p < 0.001), Escherichia-Shigella (p < 0.05) were significantly higher in gut tissue, while Romboutsia (p < 0.01) and Prevotella (p < 0.05) were significantly higher in stool samples.

The tissue biopsy samples of patients with CRC had a high abundance of bacterial of the phylum Bacteroidetes, Fusobacteria and Verrucomicrobia compared to the tissue samples obtained from healthy individuals. In contrast, Proteobacteria, Firmicutes and Actinobacteria were more abundant in tissue samples of healthy individuals compared to those with CRC (Figure 4). However, there were no significant differences of bacterial composition detected at phyla or genus level between those two sample groups. The genus Gemella was enriched in tissue of patients with CRC, while Streptococcus, Escherichia and Shigella were less abundant compared healthy controls.

**Figure 4:**
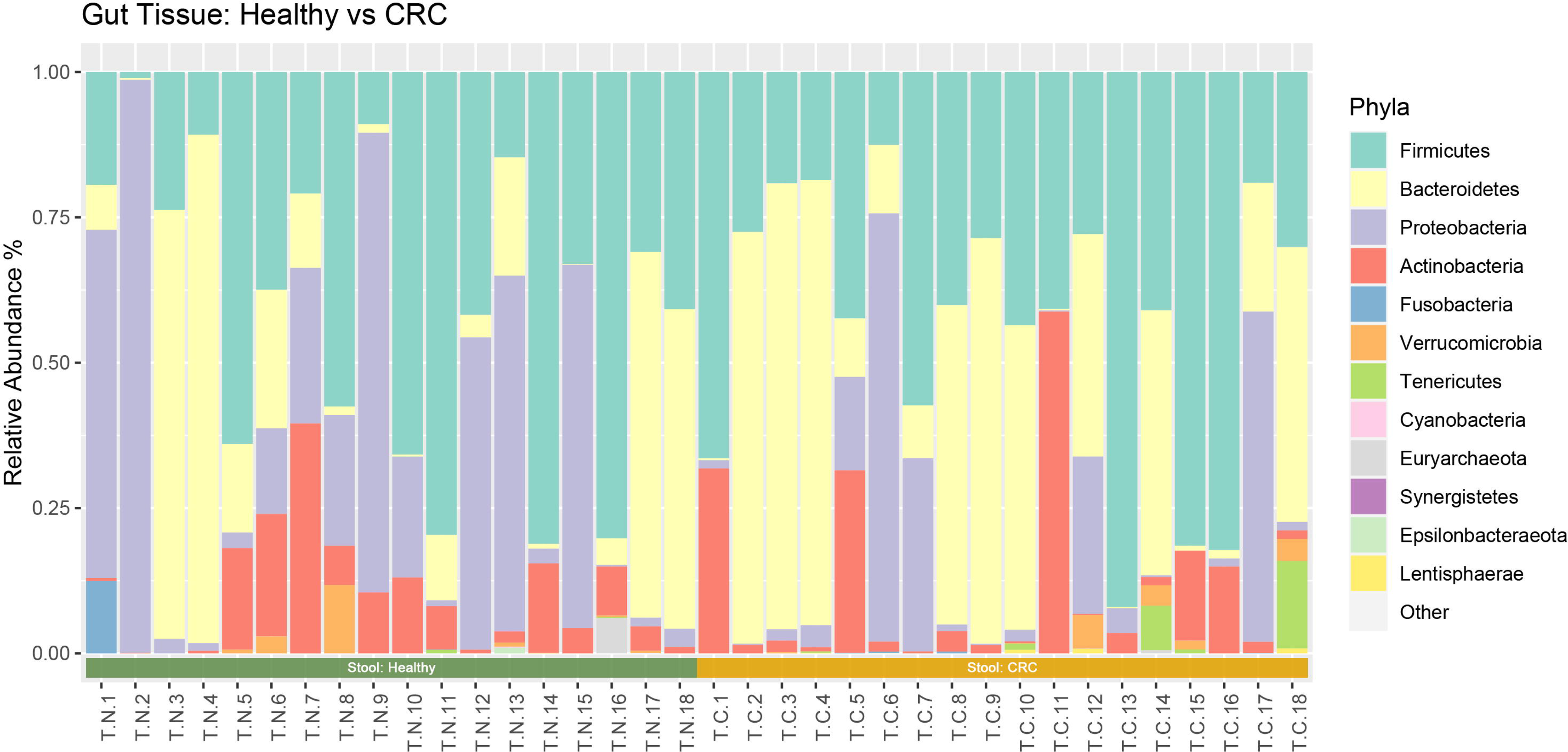
The relative abundance of Phyla of tissue samples of healthy individuals in comparison to patients with CRC. The relative abundance of different bacterial Phyla in tissue samples in healthy individuals (n=18) and in patients with CRC (n=18) was assessed. The relative abundance percentages for each sample was calculated by converting OTU counts in each sample using the R\funrar, version 1.4.1. Phylum Firmicutes, Bacteroidetes, Proteobacteria and Fusobacteria were found to be highly abundant across the tissue samples.

### Differences in the composition microbiome of stool samples in patients with pre-malignant, CRC and healthy individuals

The most abundant bacteria Phylum of stool samples of patients with CRC was Bacteroidetes compared to the stool samples of healthy and in those with pre-malignant lesions. The overall abundance of Firmicutes was higher in stool samples of those with premalignant lesions compared to healthy individuals and patients with CRC (Figure 5A). However, significant differences were only seen in the relative abundances of phylum Epsilonbacteraeota and Elusimicrobia in the stool samples of the three subgroups (healthy, those with CRC or with premalignant lesions) (p = 0.03) using the Kruskal-Wallis test (Figure 6). Post hoc pairwise testing revealed that stool samples of patients with pre-malignant lesions had significantly higher abundances (p < 0.01) of bacteria belonging to the phyla Epsilonbacteraeota and Elusimicrobia compared to healthy controls.

**Figure 5:**
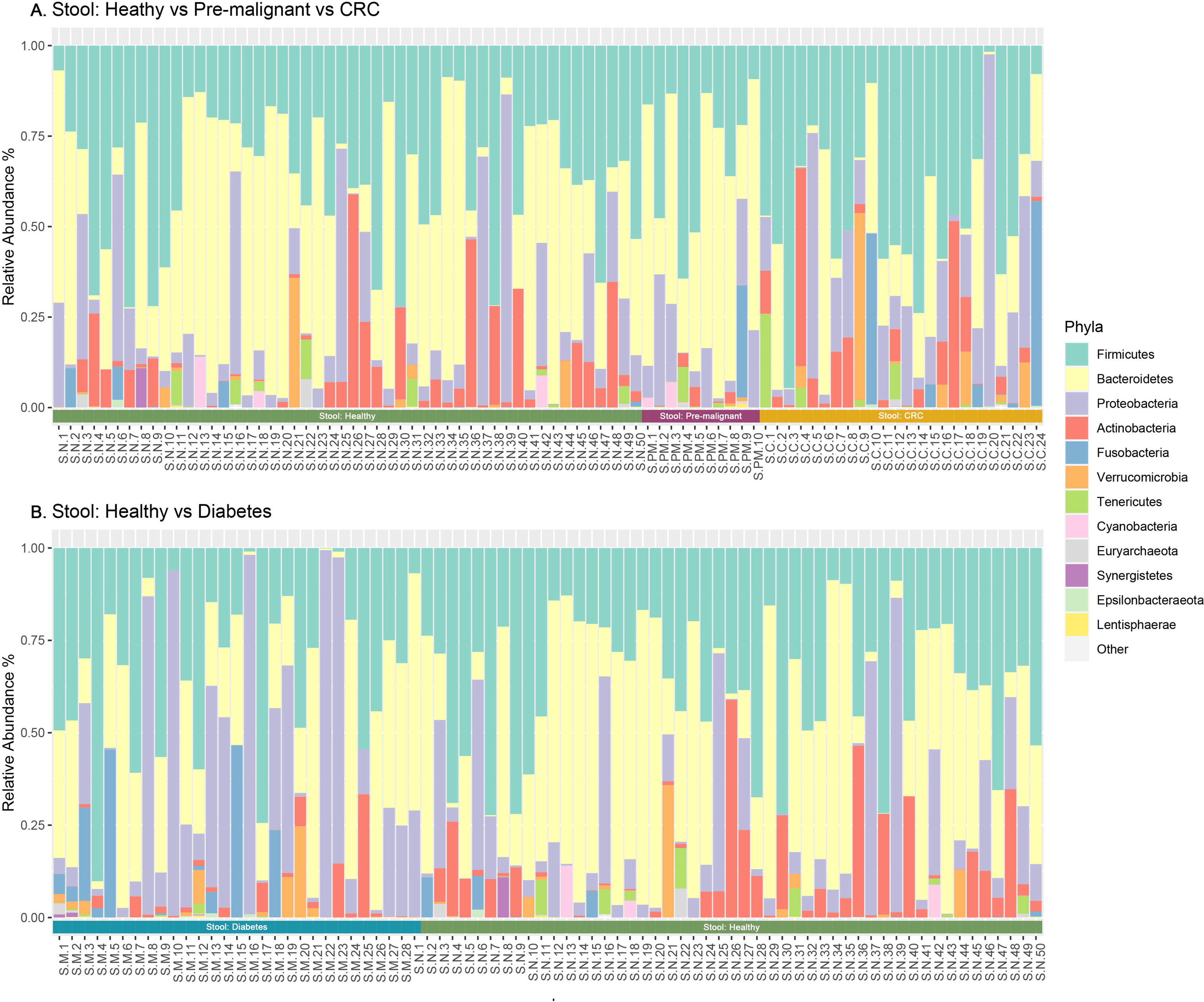
The relative abundance of phyla in stool samples of patients with CRC, healthy individuals and in those with premalignant lesions. The differences in the relative abundance of different bacterial Phyla between stool samples of healthy individuals (n=50), those with CRC (n=24), and in those with a pre-malignant lesion (n=10) were compared (A). Phylum Firmicutes, Bacteroidetes, Proteobacteria, Actinobacteria and Fusobacteria were dominant across the stool subgroups. the relative abundance of different bacterial Phyla between stool samples of healthy individuals was also compared with those of patients with diabetes (n=28) (B) Firmicutes, Bacteroidetes and Actinobacteria were found to be dominant in stool samples of individuals with diabetes. The relative abundance percentages for each sample was calculated by converting OTU counts in each sample using the R\funrar, version 1.4.1. .and healthy vs diabetes, the groups were tested at phyla and genera levels using

**Figure 6:**
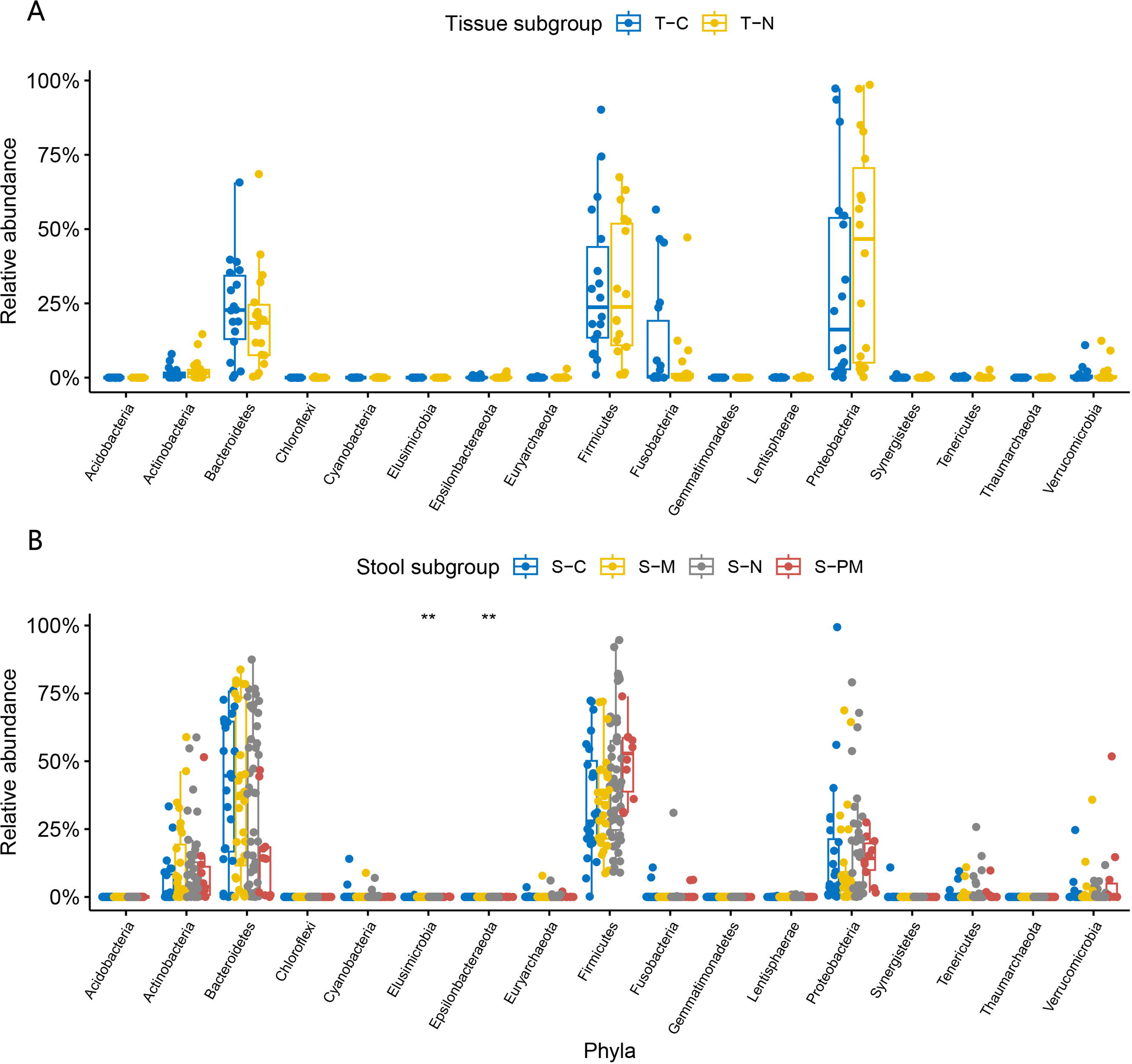
Comparison of the relative abundance of bacterial phyla in the tissue and stool samples in different subgroups. The relative abundance of bacterial phyla were compared in tissue samples (T) in healthy individuals (T-N) and in patients with CRC (T-C) using the Mann- Whiteny U test, were no difference was detected (A). The relative abundance of bacterial phyla was also compared in stool (S) samples in in those with CRC (S-C), healthy individuals (S-N), those with diabetes (S-M) and in those with premalignant lesions (S-PM), were compared using Kruskal-Wallis test with FDR corrections using Benjamini-Hochberg method (B). This showed bacteria of the Phyla of Epsilonbacteraeota and Elusimicrobia to be significantly different in stool samples between different subgroups (p = 0.03). The error bars indicate the mean and standard deviation.

There were significant differences in the relative abundance of bacteria belonging to five genera Christensenellaceae, Enterobacteriaceae, Mollicutes, Romboutsia and Ruminococcaceae in the stool samples of these three subgroups, namely healthy individuals, those with premalignant lesions and those with CRC (p < 0.05). Post-hoc tests, further confirmed that stool samples of patients with pre-malignant lesions had significantly high abundances of Christensenellaceae, Enterobacteriaceae, Mollicutes and Ruminococcaceae (p < 0.001) compared to patients with CRC, and healthy individuals. Bacteria of genus Romboutsia was significantly higher (p < 0.01) in healthy stool samples compared to the stool samples of patients with CRC.

### Differences in the composition of microbiome of stool samples in patients with diabetes mellitus and healthy individuals

As 22 (52%) of patients with CRC also had diabetes, it is not clear if the changes seen in the stool microbiome of those with CRC could be related to the presence of diabetes or specific to CRC. Therefore, we assessed the stool microbiome of 28 patients with diabetes to differentiate these observations.

Bacteria of phylum Bacteroidetes were detected at a relatively high in stool samples of patients with diabetes mellitus (38.12%) compared to healthy individuals (31.2%), while Firmicutes was found to be less abundant in patients with diabetes (35.16%) compared to healthy individuals (42.69%). Bacteria of the phylum Proteobacteria were found in similar abundance in patients with diabetes (11.81%) and in healthy individuals (13.78%) (Figure 7). At genus level, Bacteroides was enriched in stool samples of patients with diabetes (15.1%) compared to healthy individuals (9.7%) but was not significant. Genus Romboutsia was significantly depleted in patients with diabetes compared to healthy controls (p = 0.009). We did not observe any differences in relative abundance of phyla or genera the stool samples of patients with diabetes compared to patients with CRC.

**Figure 7:**
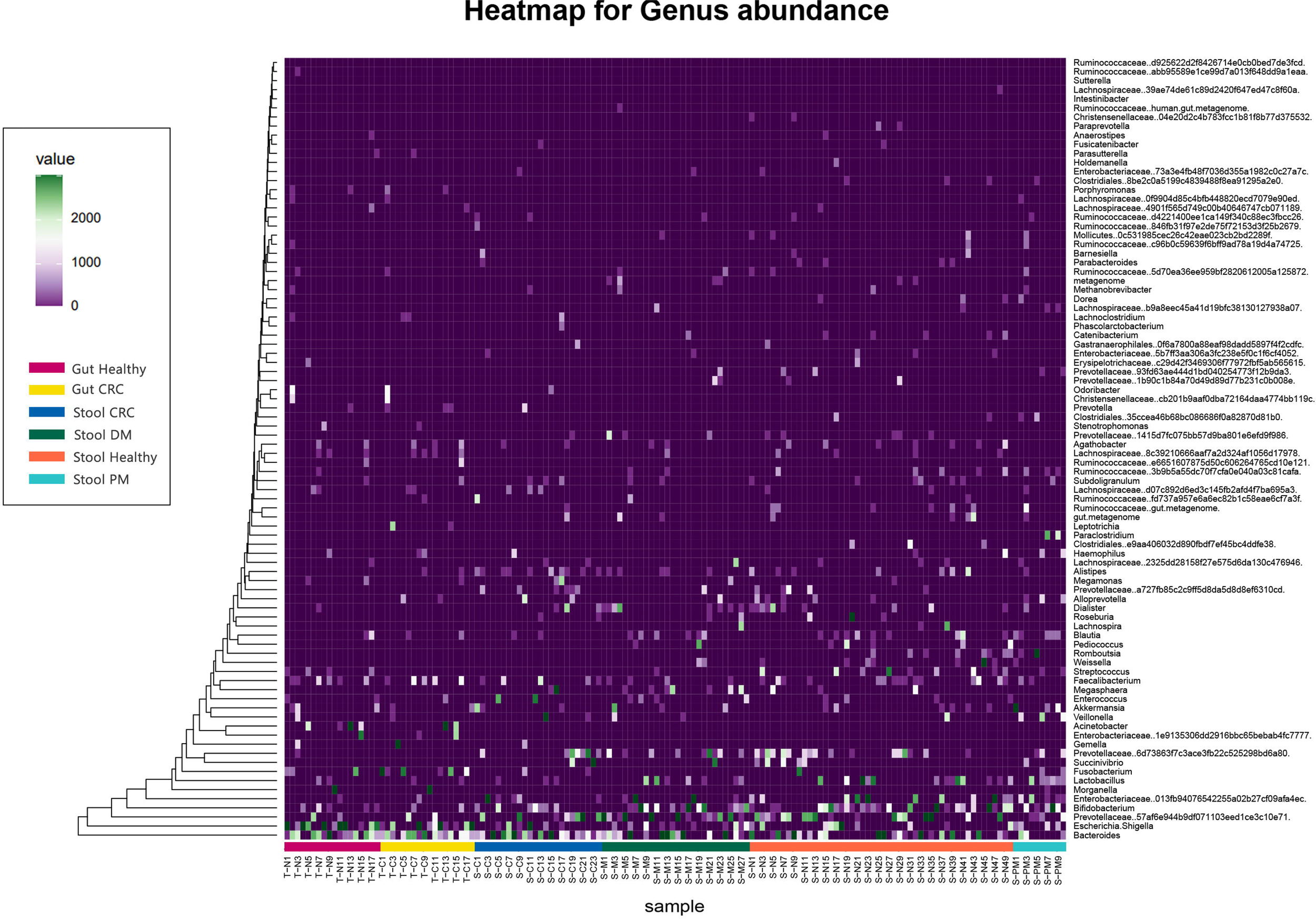
A heatmap showing relative abundance of genera in tissue and stool samples. The type of sample and the subgroup are indicated with a color key. Dendrograms were produced with the (UPGMA) method based on Bray-Curtis distance.

## Discussion

In this study first study from South Asia, we assessed the gut microbiome in stool and colonic tissue biopsies of patients with CRC and premalignant lesions, comparing them to the microbiome of healthy age and sex matched individuals and those with diabetes. Interestingly, there were significant differences in the microbiome in colonic tissue samples compared to stool samples in all subgroups, with Fusobacterium, Acinetobacter, Escherichia, Shigella being significantly more abundant in colonic tissue samples with Romboutsia and Prevotella, being the most abundant in stool samples. Although significant variations have been previously observed between stool and colonic biopsy samples, the abundance of bacteria have greatly varied ^23^ ^24^. Bacteroidetes, Fusobacteria and Verrucomicrobia were overabundant in colonic tissue samples of patients with CRC compared to healthy individuals, while Firmicutes and Actinobacteria were the most abundant in tissue samples of healthy individuals. Studies in the US and Sweden also have shown that Actinobacteria and Firmicutes were most abundant in healthy colonic tissue samples, although they did not find a high abundance of Proteobacteria^23^ ^24^. Fusobacterium have shown to be overabundant in adenomas^9^, while many studies have reported a high abundance of Bacteroides species in those with CRC compared to other groups^8^ ^9^.

Interestingly, bacteria belonging to the genus Romboutsia and Prevotella were found to be overabundant in stool samples compared to colonic tissue biopsies. Furthermore, genus Romboutsia was significantly more abundant in stool samples in healthy individuals compared to those with CRC and diabetes. Rombutsia species have shown to be less abundant in individuals with diabetes compared to healthy individuals ^25^ ^26^. However, some have shown that Rombutsia species associate with the presence of non-alcoholic fatty liver disease (NAFLD) and strongly associated with hepatocellular carcinoma (HCC), while it was less abundant in those with diabetes ^27^ ^28^. As many of these studies including ours report associations and patterns of the microbiome in disease and health, it would be important to explore if these bacteria do play a role in NAFLD and HCC and if so, the possible mechanisms involved.

In our cohort, we found that the bacteria of genus Prevotella were one of the most abundant bacteria in stool samples. *Prevotella* species have shown to associate with plant-based diets, high in fiber and low in fat content, and is highly abundant typically in individuals consuming a non-Western diet ^29^ ^30^. However, there was no difference in the abundance rates of *Prevotella* species in those with CRC, compared to healthy individuals, those with premalignant lesions or in those with diabetes. This is likely to be due to high consumption of plant-based products in the typical Sri Lankan diet. It would be important to further characterize the different bacteria at species level to understand their roles and to explore the possibility of dietary manipulation to enhance the abundance of favorable microbe species.

As shown in other studies Bacteroides species were found to be overabundant in those with CRC compared to other groups^8^ ^9^. Although we could not characterize the Bacteriodes species in this study, due to sequencing of the V4 hypervariable region of the 16S RNA, one of our previous studies using quantitative real-time PCR showed that *Bacteriodes fragilis* was significantly higher in patients with CRC compared to healthy individuals and in those with diabetes ^31^. Presence of enterotoxigenic *Bacteroides fragilis* has shown to be a potential marker for the presence of CRC and was shown to associate with poor prognosis^32^ ^33^. *Bacteroides fragilis* has shown to induce tumorigenesis by multiple mechanisms, which include alterations in NFkβ signalling pathways, inducing DNA damage, increasing polyamine metabolism, inducing TH17 cellular responses and by stimulating stem cell activity ^33^. Fusobacterium, which has also shown to associate with CRC was found to be enriched especially in colonic tissue samples in those with CRC ^10^ ^34^ ^35^. We found that those with premalignant lesions had significantly higher frequency of Christensenellaceae, Enterobacteriaceae, Mollicutes and Ruminococcaceae in their stool samples compared to patients with CRC. The presence of Christensenellaceae and Ruminococcaceae were found to be enriched in patients with adenoma previously compared to healthy individuals and have shown to be potential biomarkers for early identification of progression to CRC ^36^.

In summary, we found that despite marked differences in the Sri Lankan diet compared to the typical Western diet, *Bacteroides fragilis* and Fusobacterium species were the most abundant in those with CRC, while bacteria of genus Christensenellaceae and Ruminococcaceae were found to be most abundant in those with premalignant lesions. Interestingly, *Prevotella* species, was one of the most abundant in many individuals, possibly due to the predominant plant-based diet consumed by Sri Lankans. We believe these results pave the way for possible dietary interventions for prevention of CRC in the South Asian population.

## Acknowledgments

Funding was provided by Faculty of Medical Sciences, University of Sri Jayewardenepura. Sri Lanka. (ASP/01/RE/MED/2018/52).

## Data Availability Statement

All data is available in the manuscript, figures, and supplementary material.

## References

1. Morgan E, Arnold M, Gini A, et al. Global burden of colorectal cancer in 2020 and 2040: incidence and mortality estimates from GLOBOCAN. Gut 2023;72(2):338–44. doi: 10.1136/gutjnl-2022-327736 [published Online First: 20220908]

2. Wijeratne DT, Gunasekara S, Booth CM, et al. Colorectal Cancer Treatment Characteristics and Concordance With Guidelines in Sri Lanka: Results From a Hospital-Based Cancer Registry. JCO Glob Oncol 2022;8:e2200004. doi: 10.1200/GO.22.00004

3. Murphy CC, Zaki TA. Changing epidemiology of colorectal cancer - birth cohort effects and emerging risk factors. Nat Rev Gastroenterol Hepatol 2023 doi: 10.1038/s41575-023-00841-9 [published Online First: 20230918]

4. Kim H, Wang K, Song M, et al. A comparison of methods in estimating population attributable risk for colorectal cancer in the United States. Int J Cancer 2021;148(12):2947–53. doi: 10.1002/ijc.33489 [published Online First: 20210211]

5. Rannan-Eliya RP, Wijemunige N, Perera P, et al. Prevalence of diabetes and pre-diabetes in Sri Lanka: a new global hotspot-estimates from the Sri Lanka Health and Ageing Survey 2018/2019. BMJ Open Diabetes Res Care 2023;11(1) doi: 10.1136/bmjdrc-2022-003160

6. Le Chatelier E, Nielsen T, Qin J, et al. Richness of human gut microbiome correlates with metabolic markers. Nature 2013;500(7464):541-6. doi: 10.1038/nature12506

7. Dugas LR, Lie L, Plange-Rhule J, et al. Gut microbiota, short chain fatty acids, and obesity across the epidemiologic transition: the METS-Microbiome study protocol. BMC public health 2018;18(1):978. doi: 10.1186/s12889-018-5879-6

8. Bonnet M, Buc E, Sauvanet P, et al. Colonization of the human gut by E. coli and colorectal cancer risk. Clinical cancer research : an official journal of the American Association for Cancer Research 2014;20(4):859–67. doi: 10.1158/1078-0432.CCR-13-1343

9. Wong SH, Yu J. Gut microbiota in colorectal cancer: mechanisms of action and clinical applications. Nat Rev Gastroenterol Hepatol 2019;16(11):690–704. doi: 10.1038/s41575-019-0209-8 [published Online First: 20190925]

10. Dai Z, Coker OO, Nakatsu G, et al. Multi-cohort analysis of colorectal cancer metagenome identified altered bacteria across populations and universal bacterial markers. Microbiome 2018;6(1):70. doi: 10.1186/s40168-018-0451-2 [published Online First: 20180411]

11. Zhang FF, Cudhea F, Shan Z, et al. Preventable Cancer Burden Associated With Poor Diet in the United States. JNCI Cancer Spectr 2019;3(2):pkz034. doi: 10.1093/jncics/pkz034 [published Online First: 20190522]

12. Singh RK, Chang HW, Yan D, et al. Influence of diet on the gut microbiome and implications for human health. Journal of translational medicine 2017;15(1):73. doi: 10.1186/s12967-017-1175-y [published Online First: 20170408]

13. Leeming ER, Johnson AJ, Spector TD, et al. Effect of Diet on the Gut Microbiota: Rethinking Intervention Duration. Nutrients 2019;11(12) doi: 10.3390/nu11122862 [published Online First: 20191122]

14. Almeida A, Mitchell AL, Boland M, et al. A new genomic blueprint of the human gut microbiota. Nature 2019;568(7753):499-504. doi: 10.1038/s41586-019-0965-1 [published Online First: 20190211]

15. Dhakan DB, Maji A, Sharma AK, et al. The unique composition of Indian gut microbiome, gene catalogue, and associated fecal metabolome deciphered using multi-omics approaches. Gigascience 2019;8(3) doi: 10.1093/gigascience/giz004

16. Edgar RC. UPARSE: highly accurate OTU sequences from microbial amplicon reads. Nat Methods 2013;10(10):996–8. doi: 10.1038/nmeth.2604 [published Online First: 20130818]

17. Henderson G, Yilmaz P, Kumar S, et al. Improved taxonomic assignment of rumen bacterial 16S rRNA sequences using a revised SILVA taxonomic framework. PeerJ 2019;7:e6496. doi: 10.7717/peerj.6496 [published Online First: 20190305]

18. R: A language and environment for statistical computing [program]. Vienna, Austria: R Foundation for Statistical Computing, 2020.

19. McMurdie PJ, Holmes S. phyloseq: an R package for reproducible interactive analysis and graphics of microbiome census data. PloS one 2013;8(4):e61217. doi: 10.1371/journal.pone.0061217 [published Online First: 20130422]

20. Oksanen JB, F.G., Kindt, R., Legendre, P., Minchin, P.R., O’Hara, R.B., Simpson, G.L., Solymos, P., Stevens, M.H.H. and Wagner, H. Vegan : community ecology package version 2.4, 2015.

21. M. Grenié PD, C.M. Tucker, F. Munoz, C. Violle. funrar: An R package to characterize functional rarity. Diversity and Distributions 2017;23:1365–71.

22. Benjamini Y, Drai D, Elmer G, et al. Controlling the false discovery rate in behavior genetics research. Behavioural brain research 2001;125(1-2):279–84. doi: 10.1016/s0166-4328(01)00297-2

23. Nowicki C, Ray L, Engen P, et al. Comparison of gut microbiome composition in colonic biopsies, endoscopically-collected and at-home-collected stool samples. Front Microbiol 2023;14:1148097. doi: 10.3389/fmicb.2023.1148097 [published Online First: 20230601]

24. Vaga S, Lee S, Ji B, et al. Compositional and functional differences of the mucosal microbiota along the intestine of healthy individuals. Sci Rep 2020;10(1):14977. doi: 10.1038/s41598-020-71939-2 [published Online First: 20200911]

25. Hendricks SA, Vella CA, New DD, et al. High-Resolution Taxonomic Characterization Reveals Novel Human Microbial Strains with Potential as Risk Factors and Probiotics for Prediabetes and Type 2 Diabetes. Microorganisms 2023;11(3) doi: 10.3390/microorganisms11030758 [published Online First: 20230315]

26. Chen Z, Radjabzadeh D, Chen L, et al. Association of Insulin Resistance and Type 2 Diabetes With Gut Microbial Diversity: A Microbiome-Wide Analysis From Population Studies. JAMA Netw Open 2021;4(7):e2118811. doi: 10.1001/jamanetworkopen.2021.18811 [published Online First: 20210701]

27. Si J, Lee G, You HJ, et al. Gut microbiome signatures distinguish type 2 diabetes mellitus from non-alcoholic fatty liver disease. Comput Struct Biotechnol J 2021;19:5920–30. doi: 10.1016/j.csbj.2021.10.032 [published Online First: 20211028]

28. Feng J, Wu Y, Dai P, et al. Gut microbial signatures of patients with primary hepatocellular carcinoma and their healthy first-degree relatives. J Appl Microbiol 2023;134(10) doi: 10.1093/jambio/lxad221

29. Wu GD, Chen J, Hoffmann C, et al. Linking long-term dietary patterns with gut microbial enterotypes. Science 2011;334(6052):105-8. doi: 10.1126/science.1208344 [published Online First: 20110901]

30. Tett A, Huang KD, Asnicar F, et al. The Prevotella copri Complex Comprises Four Distinct Clades Underrepresented in Westernized Populations. Cell host & microbe 2019;26(5):666–79 e7. doi: 10.1016/j.chom.2019.08.018 [published Online First: 20191010]

31. Sahankumari A, Gamage BD, Malavige GN. Association of the gut microbiota with colorectal cancer in a South Asian cohort of patients. *bioRxiv* 2019:694125. doi: 10.1101/694125

32. Haghi F, Goli E, Mirzaei B, et al. The association between fecal enterotoxigenic B. fragilis with colorectal cancer. BMC Cancer 2019;19(1):879. doi: 10.1186/s12885-019-6115-1 [published Online First: 20190905]

33. Liu QQ, Li CM, Fu LN, et al. Enterotoxigenic Bacteroides fragilis induces the stemness in colorectal cancer via upregulating histone demethylase JMJD2B. Gut Microbes 2020;12(1):1788900. doi: 10.1080/19490976.2020.1788900 [published Online First: 20200720]

34. Farhana L, Antaki F, Murshed F, et al. Gut microbiome profiling and colorectal cancer in African Americans and Caucasian Americans. World J Gastrointest Pathophysiol 2018;9(2):47–58. doi: 10.4291/wjgp.v9.i2.47

35. Ou S, Chen H, Wang H, et al. Fusobacterium nucleatum upregulates MMP7 to promote metastasis-related characteristics of colorectal cancer cell via activating MAPK(JNK)- AP1 axis. Journal of translational medicine 2023;21(1):704. doi: 10.1186/s12967-023-04527-3 [published Online First: 20231009]

36. Wu Y, Jiao N, Zhu R, et al. Identification of microbial markers across populations in early detection of colorectal cancer. Nat Commun 2021;12(1):3063. doi: 10.1038/s41467-021-23265-y [published Online First: 20210524]

